# Self-efficacy enhanced multi-joint exercise improves fitness and cardiometabolic health in overweight and obese older adults: a randomised controlled trial

**DOI:** 10.1101/2025.10.19.25338325

**Authors:** Oratai Kanlayawut, Raweewan Maphong, Waris Wongpipit, Thanomwong Kritpet

## Abstract

Excess weight in older adults is an increasing challenge to healthy aging. While physical exercise and self-efficacy each offer benefits, few interventions combine both. This study examined the effects of integrating self-efficacy strategies with multi-joint resistance training on health outcomes in overweight and obesity older adults. Thirty-three older adults (age 67.49±4.49 years, body mass index 24.65±1.15kg/m^2^) were randomly assigned to a multi-joint exercise group (MJ) or a combined exercise and self-efficacy group (MJS). Both groups completed a 12-week progressive resistance training program (60 minutes, three times per week). The multi-joint exercise group incorporating self-efficacy theory group (MJS) also received psychological strategies based on self-efficacy theory. Outcomes included self-efficacy, body composition, physical function, and biomarkers. Both groups improved in body composition and physical function. The MJS group showed significantly greater reductions in body mass (p = 0.044), BMI (p = 0.029), and fat mass (p < 0.001), along with higher gains in self-efficacy. HbA1c levels decreased significantly in the MJS group (p = 0.021), while fasting glucose increased in the multi-joint exercise group (MJ). Combining self-efficacy strategies with resistance training enhances physical and psychological outcomes in overweight and obesity older adults and may support long-term exercise adherence.

## Introduction

The global rise in overweight and obesity among older adults presents an increasing challenge for healthcare systems. According to the World Health Organization, the age-standardized obesity rate among adults aged 18 and older grew from 6.6% in 1990 to 15.8% in 2022 [1]. These conditions contribute significantly to functional decline, reduced independence, and increased premature mortality [2, 3]. As such, addressing both the psychological and physical barriers to physical activity is essential for designing effective interventions that promote healthy aging [4].

Among these barriers, physical discomfort is a key factor deterring older adults from engaging in regular exercise. This discomfort is often linked to reduced exercise self-efficacy, the belief in one’s ability to perform physical activity, which further limits participation and worsens health outcomes [5–7]. Bandura’s self-efficacy theory identifies two major determinants of behavior change: perceived self-capability and expected outcomes. Interventions based on this theory, incorporating mastery experiences, social modeling, and verbal encouragement, have demonstrated effectiveness in boosting confidence and sustaining behavioral change [8]. Evidence supports the use of self-efficacy strategies to promote physical activity in overweight and obesity older adults, resulting in greater energy expenditure, improved exercise adherence, and better weight management [9, 10].

However, psychological readiness alone is insufficient; it must be coupled with appropriate physical training to achieve meaningful health improvements. Resistance training, particularly multi-joint exercises that engage large muscle groups through functional movements like squatting and stair climbing, has proven effective in enhancing strength, body composition, and aerobic capacity. These exercises closely mimic daily activities and offer greater functional benefits than single-joint movements, making them especially suitable for older adults [11, 12]. Even when training volume is controlled, multi-joint exercises have been shown to produce superior gains in both muscle strength and cardiovascular fitness [13]. Despite these benefits, adherence to exercise recommendations remains low among overweight and obesity older adults, often due to low self-efficacy[14].

This persistent gap underscores the need for interventions that address both the psychological and physical dimensions of behavior change. Although self-efficacy enhancement and resistance training have shown individual benefits, few studies have integrated these elements into a single, theory-based intervention. Most existing research isolates the mental or physical components, missing the opportunity to examine their synergistic effects. Therefore, the present study aims to evaluate the feasibility and effectiveness of an innovative intervention that combines self-efficacy theory with progressive multi-joint resistance training. By comparing this integrated approach to a standard resistance training program without psychological components, the study will assess the added value of incorporating self-efficacy principles. The findings may guide the development of scalable, community-based programs that foster sustainable physical and psychological well-being in overweight and obesity older adults, ultimately supporting more active, independent, and healthier aging lifestyles.

## Materials and methods

### Study design

This randomized controlled trial was conducted at the Nonthaburi City Municipality, Thailand, between March 2024 and October 2024. The study protocol was approved by the Ethical Review Committee for Research Involving Human Subjects, Health Science Group, Chulalongkorn University, Bangkok, Thailand (Approval No. 244/66). The trial was registered with the Chinese Clinical Trial Registry (ChiCTR; www.chictr.org.cn) on February 29, 2024 (Registration No. ChiCTR2400081386). All participants provided written informed consent prior to enrollment. Medical history and physical activity readiness were assessed using a standardized questionnaire. Randomization was carried out using a computerized random number generator to minimize selection bias. To ensure blinding and minimize potential bias, the sequence of experimental conditions was concealed from both investigators and participants until the beginning of each trial session. All methods were performed in accordance with the relevant guidelines and regulations.

### Sample size calculation

The sample size was calculated using G*Power 3.1.9.4 software. Based on the effect size (F = 0.4) for fat mass obtained from a previous randomized controlled trial examining the effects of resistance exercise on body composition in obese older women [15], a priori power analysis was performed for a repeated measures ANOVA, between factors. The parameters included an α error probability of 0.05, a power (1−β error probability) of 0.80, two groups, and four repeated measurements with an assumed correlation among repeated measures of 0.5. The analysis indicated that a total sample size of 34 participants would be required to detect a significant group × time interaction effect. To account for potential dropouts, 38 participants were recruited.

### Participants

Thirty-eight overweight and obese older adults (aged 60–75 years) participated in the study. Nineteen participants were randomly assigned—based on sex, age group (60–69 or 70–75 years), and body mass index (≤23–24.9 or ≥25.0 kg/m²)—to the multi-joint exercise group (MJ group), and 19 to the multi-joint exercise group incorporating self-efficacy theory (MJS group). Participants had a body mass index (BMI) ranging from 23 to 29.9 kg/m². The inclusion criteria were as follows: no history of uncontrolled diabetes, uncontrolled hypertension, cardiovascular disease, chronic obstructive pulmonary disease, or orthopedic conditions that contraindicate exercise; no use of weight-loss medication; no engagement in regular exercise or sports activities within the past three months; and low to moderate self-efficacy in performing multi-joint exercises.

### Study protocol

Venous blood samples were collected before and after the intervention, following an 8-hour overnight fast. To control for diurnal variations in blood chemistry, all samples were taken at the same time of day for both the pre-test and post-test. Two hours after breakfast, participants completed the Physical Activity Readiness Questionnaire (PAR-Q+) and a self-efficacy questionnaire specific to multi-joint exercise. Resting heart rate and blood pressure were measured while participants were seated, using a semi-automated monitor (Omron HEM-7120, Japan). Subsequently, assessments of body composition and physical function were conducted, in that order.

During the intervention phase, participants in the MJ group performed only the multi-joint exercise program, while those in the MJS group participated in the same program with the addition of self-efficacy strategies. Prior to physical function testing, all participants completed a standardized warm-up consisting of light aerobic activity and dynamic stretching exercises targeting major muscle groups. This warm-up was designed to reduce injury risk and optimize performance during assessment.

### Intervention

The multi-joint exercise group (MJ group) in 12 weeks, 3 times/week, each session lasting 60 minutes (10-min warm-up, 40-min exercise, 10-min cool-down). Each session included 8 exercises (1) shoulder press with chair squats, (2) standing row with high knees, (3) triceps extension with chair squats, (4) standing front raise with hip extension, (5) calf raise with shrugs and chair squats, (6) front lunge with lateral arm raise, (7) standing leg curls with bicep curls, and (8) split squat with bicep curls. with 2 sets of 10 repetitions (progressively increased over time) and the multi-joint exercise group incorporating self-efficacy theory (MJS group) received the same exercise program as the MJ group, with additional components based on Bandura’s self-efficacy theory strategies aimed at enhancing self-efficacy throughout the program include four primary sources: (a) mastery experience, (b) vicarious experience, (c) verbal persuasion, and (d) emotional arousal, by stimulating self-efficacy while performing multi-joint exercises simultaneously. The self-efficacy theory with multi-joint exercise protocol showed in Table 1. **Table 1** The self-efficacy theory with multi-joint exercise protocol. A 12-week program combining progressive multi-joint exercises with self-efficacy strategies to enhance exercise adherence and confidence.

**Table 1.** The self-efficacy theory with multi-joint exercise protocol. A 12-week program combining progressive multi-joint exercises with self-efficacy strategies to enhance exercise adherence and confidence.

| Week(s) | Muti-joint exercise | Self-Efficacy Theory |
| --- | --- | --- |
| 1–2 | 8 repetitions, 2 sets, 1-minute rest between sets and exercises.<br>Equipment: Women (1 kg dumbbell, 0.5 kg sandbag); Men (1.5 kg dumbbell, 1 kg sandbag) | Mastery experience: Exercise education and discussion of past exercise experiences to promote confidence. |
| 3–4 | 10 repetitions, same rest interval and sets. | Mastery experience (continued): Reinforcing skills and promoting belief in exercise capability. |
| 5–6 | Increased resistance:<br>Women (1.5 kg dumbbell, 1 kg sandbag);<br>Men (2 kg dumbbell, 1.5 kg sandbag) | Mastery experience and guided sessions:<br>Instructor-led activities with researcher guidance and feedback. |
| 7–8 | Increased to 3 sets, same resistance. | Mastery and vicarious experience:<br>Introduction to role modeling by peers. |
| 9–12 | Increased resistance:<br>Women (1.5 kg dumbbell, 1.5 kg sandbag);<br>Men (2 kg dumbbell, 2 kg sandbag) | Vicarious experience and self-modeling:<br>Peer leadership and participant role models to reinforce efficacy. |
| 1–12 | Sessions were supervised by an external instructor with researcher support, while participants engaged in self-monitoring through exercise diaries. Verbal persuasion and emotional arousal were facilitated via encouragement, motivational messages, and observation of affective responses. |  |
| 5–12 | Behavioral reinforcement was provided through structured praise and social rewards recognizing leadership and effort. |  |
| 6–12 | Peer-led sessions facilitated vicarious experience through observation and interaction with successful role models. |  |

### Measurement

The outcome measures, which focused on evaluating self-efficacy in muti-joint exercise, body composition, physical function, and blood chemistry, were administered at pre-test (week 0) and post-test (week 12).

### Self-efficacy in multi-joint exercise questionnaire

The reliability of a self-efficacy capability questionnaire for designing multi-joint exercise programs in overweight and obesity older adults. Tested on 40 participants, the questionnaire showed high internal consistency with a Cronbach’s Alpha of 0.90. Subscale scores were 0.72 for knowledge, 0.88 for self-efficacy capability, and 0.76 for expected outcomes, confirming the questionnaire’s reliability for assessing exercise planning capability. The assessment criteria are based on an adapted criterion-referenced approach, following Bloom’s principles and evaluation methods[16]. The questionnaire consists of 4 parts were as follows: The first part gathered general and health information, including name, age, underlying diseases, and self-care through exercise.

The second part assessed knowledge about multi-joint exercises using a multiple-choice format, with a total of 10 questions. Each question had two answer choices: “Yes” and “No.” The scoring criteria are as follows: answering “Yes” earns 1 point, while answering “No” earns 0 points. A low level is 0–4.9, a medium level is 5–7.9, and a high level is 8–10.

The third part assessed self-efficacy in multi-joint exercises, and the fourth part assessed perceived outcomes of multiple-choice exercises, both using a multiple-choice format with a total of 10 questions. Each question had three answer choices: “Yes,” “Unsure,” and “No.” The questions included both positive and negative statements. The scoring criteria are as follows: for positive statements, answering “Yes” earns 3 points, “Unsure” earns 2 points, and “No” earns 1 point; for negative statements, the scoring is reversed: “Yes” earns 1 point, “Unsure” earns 2 points, and “No” earns 3 points. The scoring range is 10–30 points, categorized into three levels: a low level (10–16), a medium level (17–24), and a high level (25–30).

### Body composition

Body composition was evaluated using a bioelectrical impedance analyzer (BIA), specifically the Omron HBF-375, Japan measuring parameters such as percentage body fat, fat mass, fat-free mass, and muscle mass.

### Physical function

A set of physical function measures followed the guidelines from the Senior Fitness Test[17].

### Muscle strength

Muscle strength testing was conducted for both the upper and lower body using a back and leg dynamometer, Takei Scientific Instruments, Takei Back and Leg Dynamometer T.K.K.5002, Japan as well as a hand grip dynamometer, Takei Scientific Instruments, Takei Hand Grip Dynamometer T.K.K.5401, Japan) Each test was performed twice, and the best value was recorded.

### Muscular endurance

Muscular endurance testing conducted for both the upper and lower body using the 30-sec arm curl test dumbbells weighing 2 kg for men and 1 kg for women and the 30-sec chair stand test. The number of repetitions performed within the given time was recorded.

### Balance testing

Balance testing was conducted for both static and dynamic balance using the single-leg stance test and the Timed Up and Go test. The recorded values were measured in seconds.

### Blood Collection and Biochemical Analysis

Venous blood samples were collected by a certified medical technologist from the antecubital vein after an overnight fasting period of at least 8 hours. Blood for glucose and lipid profile analyses was collected into serum separator tubes, while blood for HbA1c analysis was collected into dipotassium ethylenediaminetetraacetic acid (K_2_EDTA) tubes All biochemical analyses were performed immediately following collection at a certified clinical laboratory within the Faculty of Allied Health Sciences, Chulalongkorn University, Pathumwan, Thailand.

Fasting plasma glucose (FPG) and lipid profiles (total cholesterol, triglycerides, high-density lipoprotein cholesterol (HDL-c), and low-density lipoprotein cholesterol (LDL-c)) were analyzed using an automated chemistry analyzer (Beckman Coulter AU480, USA) employing enzymatic colorimetric methods. The intra-assay coefficient of variation (CV) for glucose and lipid measurements was less than 3%. Glycated hemoglobin (HbA1c) was measured using high-performance liquid chromatography (HPLC) on a Bio-Rad D-10™ Hemoglobin Testing System (Bio-Rad Laboratories, USA). The intra-assay CV for HbA1c measurement was below 2%.

High-sensitivity C-reactive protein (hs-CRP) was assessed via an immunoturbidimetric assay using the same chemistry analyzer (Beckman Coulter AU480). The intra-assay CV for hs-CRP was less than 5%. All assays were performed according to the manufacturers’ protocols and quality control standards. Blood samples were processed and analyzed within two hours of collection to ensure data accuracy.

### Statistical analyzes

Statistical analyses were conducted using SPSS (version 26; IBM Corp., Armonk, NY, USA). The normality and homogeneity of variance for each variable were assessed using the Shapiro– Wilk test and Levene’s test, respectively, prior to further statistical analysis. Body composition, physical function, blood chemistry, and self-efficacy in multiple-joint exercise were analyzed using a two-way ANOVA (group × time) with repeated measures. For multiple comparisons, Fisher’s least significant difference (LSD) test was used as a post hoc analysis. For variables that violated the assumption of normality, non-parametric comparisons between groups were conducted using the Mann–Whitney U test. Statistical significance was set at a two-tailed p-value of < 0.05. Effect sizes were calculated to assess the magnitude of the observed differences. For parametric tests, effect sizes were reported as partial eta squared (η²p), depending on the type of analysis. For analyses of variance (e.g., ANOVA), partial eta squared (η²p) was reported and interpreted as: small (0.01), medium (0.06), and large (0.14). For non-parametric tests, effect sizes were interpreted using the rank-biserial correlation or similar non-parametric measures, with the following thresholds: small (< 0.1), medium (∼0.3), and large (> 0.5) [18, 19].

## Results

Five participants dropped out during the study due to time constraints (two from the MJS group and three from the MJ group). Table 2. presents the general physiological characteristics and body composition of both groups. After the intervention, resting heart rate (HR) significantly increased in the MJ group (68.31±10.35 vs. 75.75±10.51; p **=** 0.001) but showed a slight, non-significant increase in the MJS group. Systolic blood pressure (SBP) slightly decreased in the MJS group but increased in the MJ group. Both groups experienced reductions in body mass, BMI, and fat mass. The MJS group showed significant decreases in body mass (59.47±5.49 vs. 57.85±4.84; p <0.001), BMI (24.56±1.21 vs. 23.82±1.42; p <0.001), and fat mass (20.84±2.50 vs. 19.59±2.58; p <0.001), whereas the MJ group had a significant reduction only in fat mass (22.23±2.40 vs. 21.43±2.44; p = 0.012). Fat-free mass and muscle mass remained largely unchanged in both groups. In the post-test, fat mass was significantly lower in MJS group compared to MJ group (19.59±2.58 vs. 21.43±2.44; p <0.001). A significant interaction effect (group × time) was found for resting HR (p = 0.026), body mass (p = 0.044), BMI (p = 0.029), and fat mass (p < 0.001), but not for fat-free mass or muscle mass.

**Table 2.**
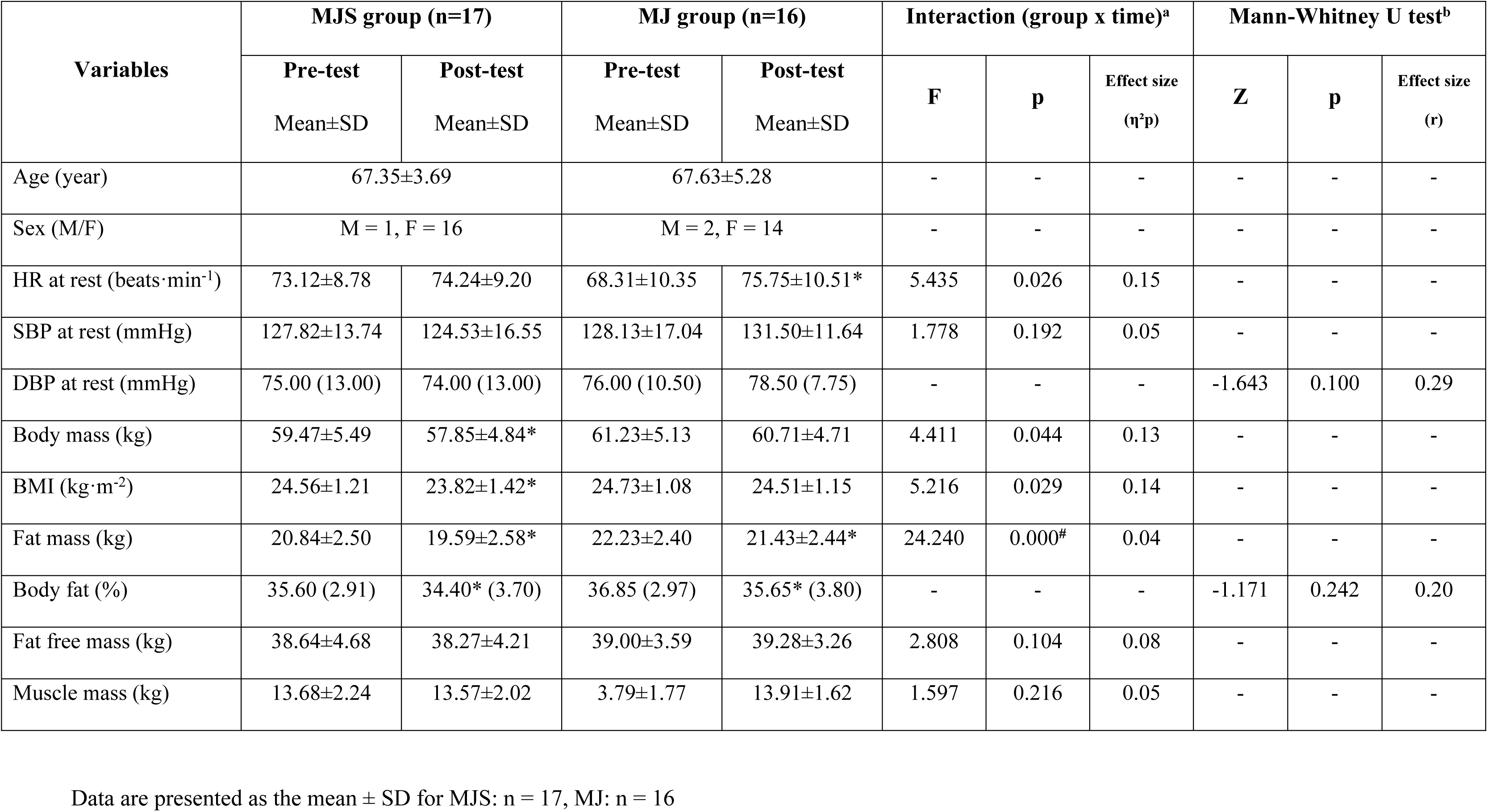
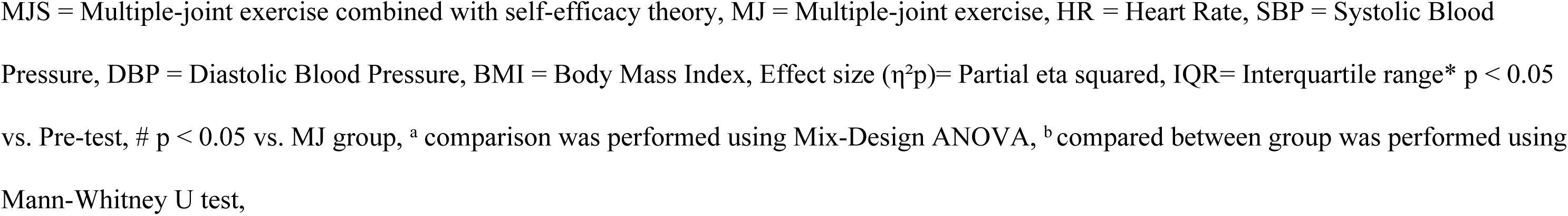
General Physiological Characteristics and Body composition of Multiple-joint exercise combined with self-efficacy theory and Multiple-joint exercise groups. Data are presented as the mean ± SD for MJS: n = 17, MJ: n = 16. * p ˂ 0.05 vs. Pre-test, # p ˂ 0.05 vs. MJ group.

| Variables | MJS group (n=17) |  | MJ group (n=16) |  | Interaction (group x time)<sup>a</sup> |  |  | Mann-Whitney U test<sup>b</sup> |  |  |
| --- | --- | --- | --- | --- | --- | --- | --- | --- | --- | --- |
| | Pre-test<br>Mean±SD | Post-test<br>Mean±SD | Pre-test<br>Mean±SD | Post-test<br>Mean±SD | F | p | Effect size<br>($\eta^2p$) | Z | p | Effect size<br>(r) |
| Age (year) | 67.35±3.69 |  | 67.63±5.28 |  | - | - | - | - | - | - |
| Sex (M/F) | M = 1, F = 16 |  | M = 2, F = 14 |  | - | - | - | - | - | - |
| HR at rest (beats·min<sup>-1</sup>) | 73.12±8.78 | 74.24±9.20 | 68.31±10.35 | 75.75±10.51* | 5.435 | 0.026 | 0.15 | - | - | - |
| SBP at rest (mmHg) | 127.82±13.74 | 124.53±16.55 | 128.13±17.04 | 131.50±11.64 | 1.778 | 0.192 | 0.05 | - | - | - |
| DBP at rest (mmHg) | 75.00 (13.00) | 74.00 (13.00) | 76.00 (10.50) | 78.50 (7.75) | - | - | - | -1.643 | 0.100 | 0.29 |
| Body mass (kg) | 59.47±5.49 | 57.85±4.84* | 61.23±5.13 | 60.71±4.71 | 4.411 | 0.044 | 0.13 | - | - | - |
| BMI (kg·m<sup>-2</sup>) | 24.56±1.21 | 23.82±1.42* | 24.73±1.08 | 24.51±1.15 | 5.216 | 0.029 | 0.14 | - | - | - |
| Fat mass (kg) | 20.84±2.50 | 19.59±2.58* | 22.23±2.40 | 21.43±2.44* | 24.240 | 0.000<sup>#</sup> | 0.04 | - | - | - |
| Body fat (%) | 35.60 (2.91) | 34.40* (3.70) | 36.85 (2.97) | 35.65* (3.80) | - | - | - | -1.171 | 0.242 | 0.20 |
| Fat free mass (kg) | 38.64±4.68 | 38.27±4.21 | 39.00±3.59 | 39.28±3.26 | 2.808 | 0.104 | 0.08 | - | - | - |
| Muscle mass (kg) | 13.68±2.24 | 13.57±2.02 | 3.79±1.77 | 13.91±1.62 | 1.597 | 0.216 | 0.05 | - | - | - |

MJS = Multiple-joint exercise combined with self-efficacy theory, MJ = Multiple-joint exercise, HR = Heart Rate, SBP = Systolic Blood Pressure, DBP = Diastolic Blood Pressure, BMI = Body Mass Index, Effect size ( $\eta^2p$ )= Partial eta squared, IQR= Interquartile range\* $p < 0.05$ vs. Pre-test, # $p < 0.05$ vs. MJ group, <sup>a</sup> comparison was performed using Mix-Design ANOVA, <sup>b</sup> compared between group was performed using Mann-Whitney U test,

Table 3. presents the physical function outcomes of both groups. After the intervention, both groups showed significant improvements in several measures. The MJ group showed a significant increase in both right- and left-hand grip strength (23.56±3.83 vs. 25.69±3.74, 22.09±4.52 vs. 23.94±4.88, p=0.007, p = 0.003 respectively), while the MJS group exhibited significant improvements in left-hand grip strength (22.71±4.77 vs. 24.41±5.28; p = 0.005). Back strength significantly improved in both groups (45.47±13.04 vs. 54.65±13.68, 43.38±9.77 vs. 56.63±12.84; all ps < 0.05, respectively), whereas leg strength increased in both groups but did not reach statistical significance. The 30-second arm curl test showed significant improvements in both right and left arms for both the MJS and MJ groups (right arm: 21.12±5.61 vs. 32.71±6.85, 19.44±6.29 vs. 29.19±6.24; left arm: 22.71±5.54 vs. 33.94±7.50, 19.06±6.18 vs. 30.25±7.17; p < 0.001 for all comparisons). Similarly, the 30-second chair stand test improved in both the MJS and MJ groups, but the increase was not statistically significant. Balance, measured by the single-leg stance test, improved significantly in the MJ group (21.25 vs. 28.38; p = 0.039), whereas the MJS group showed a slight but non-significant improvement. Balance testing demonstrated a significant reduction in completion time in both the MJS and MJ groups (7.85±1.10 vs. 6.41±0.69, 7.96±0.85 vs. 6.63±1.20; p < 0.001 for all comparisons), indicating enhanced mobility. Despite these improvements, interaction effects (group × time) were not significant for any measure (p > 0.05), suggesting similar progress in both groups.

**Table 3.**
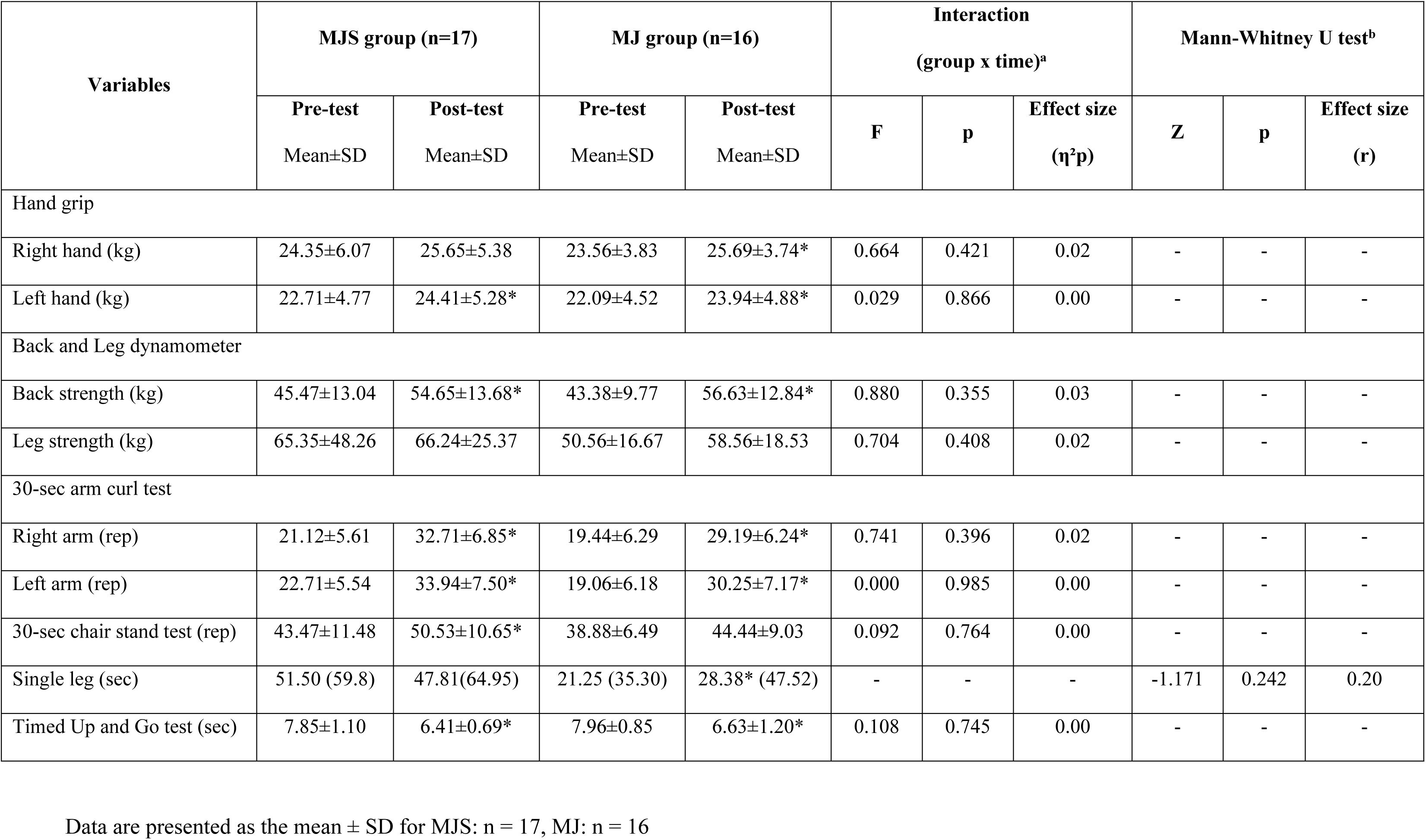
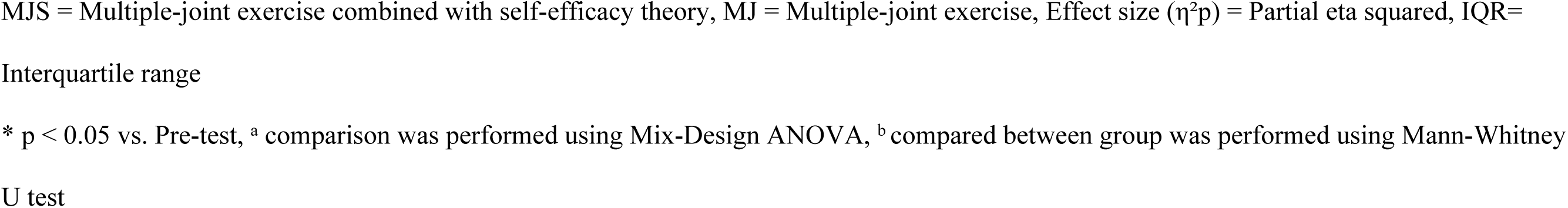
Physical function of Multiple-joint exercise combined with self-efficacy theory and Multiple-joint exercise groups. Data are presented as the mean ± SD for MJS: n = 17, MJ: n = 16. * p ˂ 0.05 vs. Pre-test

| Variables | MJS group (n=17) |  | MJ group (n=16) |  | Interaction<br>(group x time)<sup>a</sup> |  |  | Mann-Whitney U test<sup>b</sup> |  |  |
| --- | --- | --- | --- | --- | --- | --- | --- | --- | --- | --- |
| | Pre-test<br>Mean±SD | Post-test<br>Mean±SD | Pre-test<br>Mean±SD | Post-test<br>Mean±SD | F | p | Effect size<br>($\eta^2p$) | Z | p | Effect size<br>(r) |
| Hand grip |  |  |  |  |  |  |  |  |  |  |
| Right hand (kg) | 24.35±6.07 | 25.65±5.38 | 23.56±3.83 | 25.69±3.74* | 0.664 | 0.421 | 0.02 | - | - | - |
| Left hand (kg) | 22.71±4.77 | 24.41±5.28* | 22.09±4.52 | 23.94±4.88* | 0.029 | 0.866 | 0.00 | - | - | - |
| Back and Leg dynamometer |  |  |  |  |  |  |  |  |  |  |
| Back strength (kg) | 45.47±13.04 | 54.65±13.68* | 43.38±9.77 | 56.63±12.84* | 0.880 | 0.355 | 0.03 | - | - | - |
| Leg strength (kg) | 65.35±48.26 | 66.24±25.37 | 50.56±16.67 | 58.56±18.53 | 0.704 | 0.408 | 0.02 | - | - | - |
| 30-sec arm curl test |  |  |  |  |  |  |  |  |  |  |
| Right arm (rep) | 21.12±5.61 | 32.71±6.85* | 19.44±6.29 | 29.19±6.24* | 0.741 | 0.396 | 0.02 | - | - | - |
| Left arm (rep) | 22.71±5.54 | 33.94±7.50* | 19.06±6.18 | 30.25±7.17* | 0.000 | 0.985 | 0.00 | - | - | - |
| 30-sec chair stand test (rep) | 43.47±11.48 | 50.53±10.65* | 38.88±6.49 | 44.44±9.03 | 0.092 | 0.764 | 0.00 | - | - | - |
| Single leg (sec) | 51.50 (59.8) | 47.81(64.95) | 21.25 (35.30) | 28.38* (47.52) | - | - | - | -1.171 | 0.242 | 0.20 |
| Timed Up and Go test (sec) | 7.85±1.10 | 6.41±0.69* | 7.96±0.85 | 6.63±1.20* | 0.108 | 0.745 | 0.00 | - | - | - |

MJS = Multiple-joint exercise combined with self-efficacy theory, MJ = Multiple-joint exercise, Effect size ( $\eta^2p$ ) = Partial eta squared, IQR= Interquartile range \* $p < 0.05$ vs. Pre-test, <sup>a</sup> comparison was performed using Mix-Design ANOVA, <sup>b</sup> compared between group was performed using Mann-Whitney U test

The MJS group demonstrated significant reductions in total cholesterol (5.97 ± 1.57 to 5.38 ± 1.41 mmol/L), HDL-c (1.49 ± 0.29 to 1.39 ± 0.21 mmol/L), and HbA1c levels (37.90 ± 4.92 to 36.87 ± 4.79 mmol/mol) following the intervention (p =0.022, p = 0.034, p < 0.001, respectively). However, a significant increase in triglyceride levels was observed compared to pre-test values. In contrast, the MJ group exhibited a significant increase in triglyceride and fasting plasma glucose (FPG) (1.13±0.35 vs. 1.44±0.44; 5.20±0.63 vs. 5.46±0.51; p = 0.001, p = 0.002, respectively). After the intervention HbA1c levels were significantly lower in the MJS group compared to the MJ group (36.87±4.79 vs. 38.87±5.54; p = 0.021). A significant interaction effect (group × time) was found for HbA1c (p = 0.021).

Moreover, hs-CRP tended to decrease in the MJS group, while it increased in the MJ group (1.14 vs. 0.81; 1.19 vs. 1.20, respectively) (In Table 4).

**Table 4.** Blood Chemistry of Multiple-joint exercise combined with self-efficacy theory and Multiple-joint exercise groups. Data are presented as the mean ± SD for MJS: n = 17, MJ: n = 16. * p ˂ 0.05 vs. Pre-test, # p ˂ 0.05 vs. MJ group.

| Variables | MJS group (n=17) |  | MJ group (n=16) |  | Interaction (group x time)<sup>a</sup> |  |  | Mann-Whitney U test<sup>b</sup> |  |  |
| --- | --- | --- | --- | --- | --- | --- | --- | --- | --- | --- |
| | Pre-test<br>Mean±SD | Post-test<br>Mean±SD | Pre-test<br>Mean±SD | Post-test<br>Mean±SD | F | p | Effect size<br>($\eta^2p$) | Z | p | Effect size<br>(r) |
| Total cholesterol<br>(mmol/L) | 5.97±1.57 | 5.38±1.41* | 5.43±1.03 | 5.52±0.92 | 3.650 | 0.065 | 0.11 | - | - | - |
| Triglyceride<br>(mmol/L) | 1.17±0.39 | 1.37±0.51* | 1.13±0.35 | 1.44±0.44* | 0.738 | 0.397 | 0.02 | - | - | - |
| HDL-c (mmol/L) | 1.49±0.29 | 1.39±0.21* | 1.51±0.27 | 1.49±0.29 | 1.644 | 0.210 | 0.05 | - | - | - |
| LDL-c (mmol/L) | 3.01±0.72 | 2.83±0.43 | 3.32±0.74 | 3.22±0.40 | 0.099 | 0.756 | 0.00 | - | - | - |
| FPG (mmol/L) | 4.98±0.44 | 5.04±0.36 | 5.20±0.63 | 5.46±0.51* | 3.059 | 0.091 | 0.10 | - | - | - |
| HbA1C (mmol) | 37.90±4.92 | 36.87±4.79* | 39.00±4.87 | 38.87±5.54 | 5.925 | 0.021<sup>#</sup> | 0.16 | - | - | - |
| hs-CRP (mg/L) | 1.14 (1.68) | 0.81 (0.77) | 1.19 (1.17) | 1.20 (2.1) | - | - | - | -1.495 | 0.135 | 0.21 |

MJS = Multiple-joint exercise combined with self-efficacy theory, MJ = Multiple-joint exercise, HDL-c = HD, LDL-c = Low-density
Lipoprotein, FPG = Fasting plasma glucose, HbA1C = Hemoglobin A1C, mmol/L = (mg/dL)/Conversion factor, Effect size ( $\eta^2p$ ) = Partial eta
squared, IQR= Interquartile range \* p < 0.05 vs. Pre-test, # p < 0.05 vs. MJ group, <sup>a</sup> comparison was performed using Mix-Design ANOVA, <sup>b</sup>
compared between group was performed using Mann-Whitney U test.

Finally, Fig 2 illustrates changes in self-efficacy component knowledge, self-efficacy, and expectation between the MJS group and the MJ group. The MJS group demonstrated a significant increase in knowledge scores from pre-test to post-test (5.91±1.76 vs. 8.13±1.32; p < 0.001), with post-test values also significantly higher than those of the MJ group (5.91p±1.76 vs. 7.55±1.74; p < 0.05). The MJ group showed no significant change. Self-efficacy scores significantly improved in both groups from pre-test to post-test (MJS: 20.74±3.15 vs. 29.13±1.46; MJ: 21.45±1.99 vs. 28.09±2.24; p < 0.001, p = 0.001, respectively); however, the MJS group exhibited a significantly greater increase compared to the MJ group (29.13±1.46 vs. 28.09±2.24; p < 0.001). Expectation scores in the MJS group significantly increased from pre-test to post-test (27.22±3.38 vs. 29.04±1.33; p = 0.009) and were also significantly higher than those of the MJ group after the intervention (29.04±1.33 vs 27.86±2.00; p < 0.001). No significant change was observed in the MJ group.

**Fig 1.**
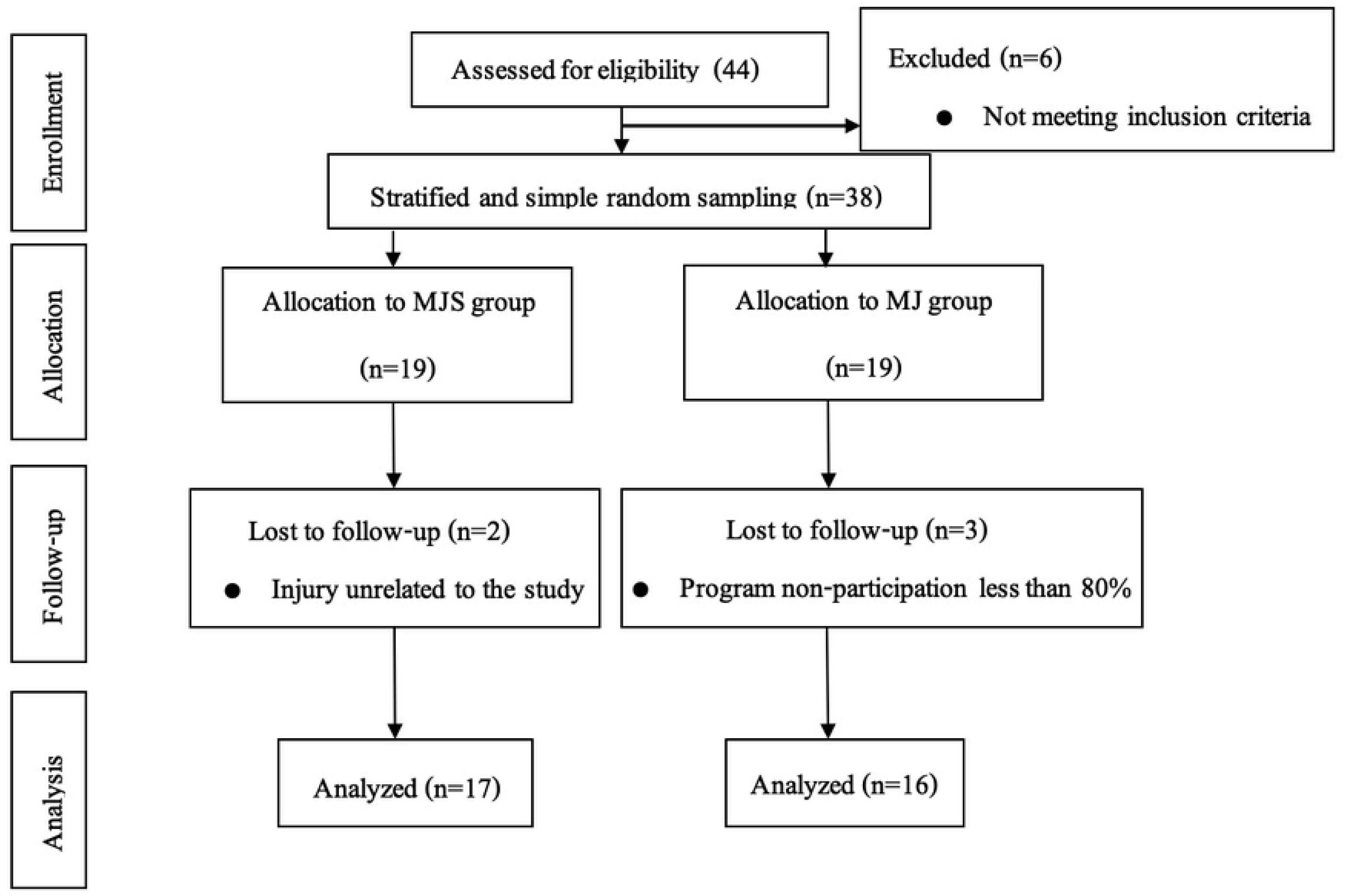
CONSORT flow diagram showing participant enrollment. A total of 44 older adults were screened for eligibility; 38 were randomized into the multi-joint self-efficacy (MJS) group (n=19) and multi-joint (MJ) group (n=19). After follow-up losses, 17 participants in the MJS group and 16 in the MJ group were included in the final analysis.

**Fig 2.**
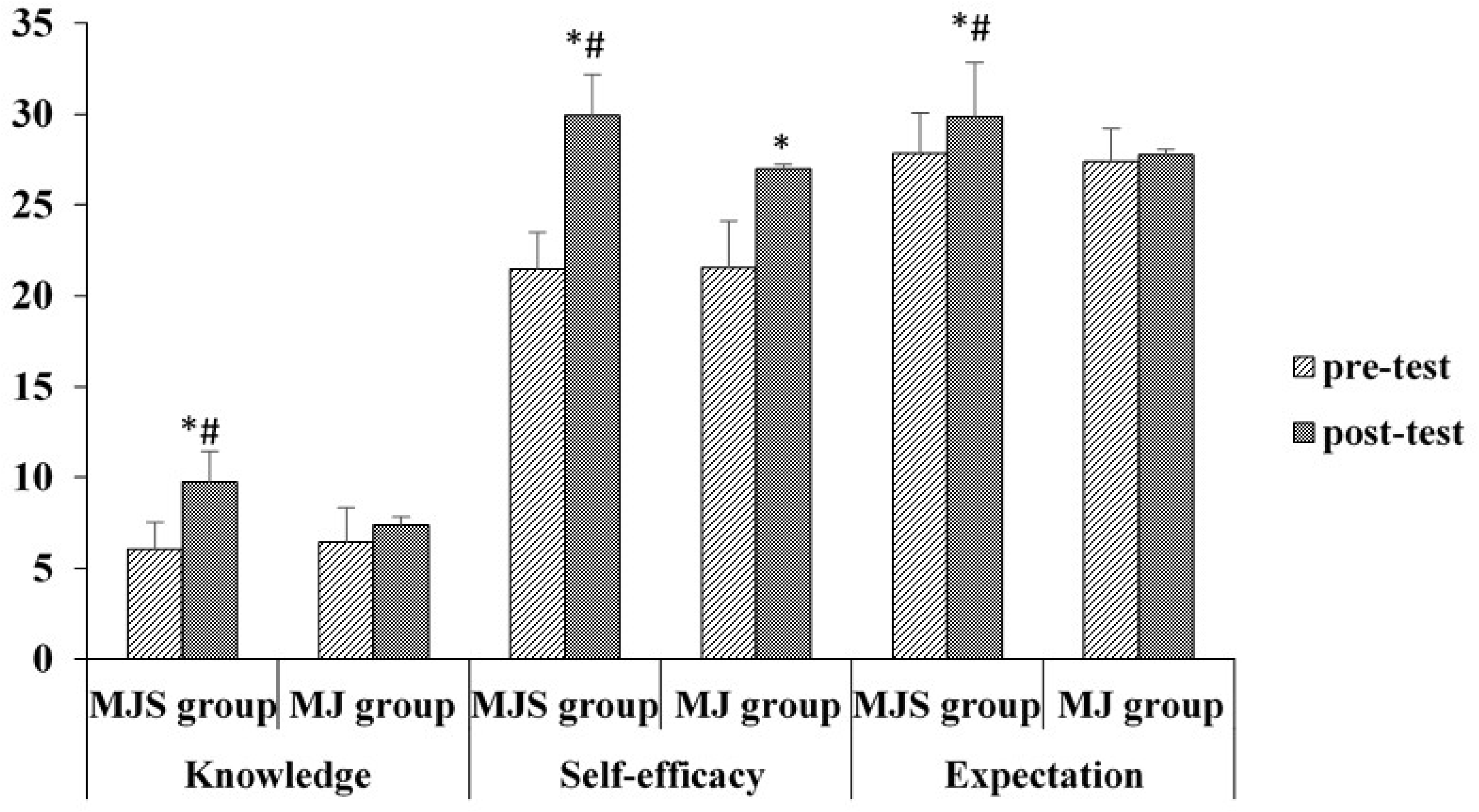
Score of self-efficacies in multiple-joint exercise. Pre- and post-test comparisons of knowledge, self-efficacy, and outcome expectation between the multi-joint self-efficacy (MJS) and multi-joint (MJ) groups. *p < 0.05 vs pre-test; #p < 0.05 vs MJ group.

## Discussion

The results of this study demonstrate the efficacy of combining multiple-joint exercise with self-efficacy theory (MJS group) in improving body composition, physical function, and glycemic control in older adults. Compared to multiple-joint exercise alone (MJ group), the integration of self-efficacy theory appears to provide additional benefits, particularly in reducing fat mass and improving blood glucose regulation. Furthermore, the findings support the notion that MJS-based interventions are more effective than general MJ programs in enhancing participants’ knowledge, self-efficacy, and expectations.

The significant reduction in body mass and BMI observed in the MJS group, along with a decrease in fat mass in both groups, aligns with previous research emphasizing the positive effects of multi-joint exercises resistance training on body composition[13, 20]. However, the MJS group exhibited greater reductions in fat mass, suggesting that self-efficacy theory may enhance adherence, motivation, and exercise intensity, leading to improved outcomes. Previous studies have shown that improvements in exercise self-efficacy significantly influence the enhancement of health-promoting behaviors. Specifically, increases in exercise self-efficacy indirectly contribute to initial weight loss by promoting healthier behaviors. Additionally, the rise in health-promoting behaviors has a significant impact on initial weight loss as well [21].

Moreover, greater frequencies of physical activity (PA) were linked to higher exercise self-efficacy, highlighting its critical role in the relationship between physical activity and weight management [22]. Therefore, integrating multi-joint exercises with self-efficacy theory may provide a synergistic approach to maintaining body composition. This combined strategy appears to be more effective than implementing exercise or self-efficacy theory alone in supporting weight loss among overweight and obesity older adults.

Both groups showed notable improvements in physical function, including handgrip strength, arm curl repetitions, and chair stand performance, which are crucial for maintaining independence and quality of life in older adults. Interestingly, the MJ group demonstrated superior performance in the single-leg balance test. This may indicate that balance-specific improvements could be more influenced by task-specific training rather than psychological interventions. Previous studies have compared task-specific training with resistance training in older adults and found that both groups showed improvements in balance. However, the group that underwent task-specific training achieved superior outcomes [23]. Other studies have also indicated that task-specific balance training improves physical function, reduces pain, enhances daily activities, and improves quality of life in patients with balance impairments [24]. Therefore, task-specific training may be more effective than psychological interventions in improving balance in overweight and obesity older adults.

Additionally, improvements in the balance testing highlight the effectiveness of both exercise interventions in enhancing functional mobility and reducing fall risk. The effectiveness of a combined exercise intervention, incorporating both resistance and balance training, has been demonstrated to improve the balance testing performance, a key indicator of fall risk reduction in older adults [11]. An eight-week resistance training program has also been shown to enhance the balance testing performance and self-efficacy scores, both of which are negatively associated with fear of falling in older adults [12]. Therefore, improvements in the balance testing performance may be attributed to resistance training and high self-efficacy, both of which are equally important for overweight and obesity older adults.

The reductions in HbA1C in both groups and the stable FPG levels in the MJS group are significant findings. The greater glycemic control observed in the MJS group suggests that self-efficacy-enhanced exercise promotes better metabolic outcomes. An improvement in self-efficacy has been linked to a higher likelihood of maintaining normal HbA1C levels in older adults [25]. The enhancement of self-efficacy through theory-based interventions can effectively improve self-efficacy, leading to better self-care activities and reduced HbA1C levels [26]. These results align with evidence suggesting that psychological interventions may improve adherence to healthy behaviors, thereby influencing glucose metabolism. Several diabetes-specific psychological factors may contribute to the relationship between psychological distress and glycemic control [27]. Moreover, hs-CRP decreased in the MJS group, while it increased in the MJ group. Typically, the reduction in inflammation (hs-CRP) may be attributed to resistance training, which enhances blood circulation, increases muscle mass, and reduces fat tissue. These changes are associated with a decrease in inflammatory markers (hs-CRP) [28, 29]. However, in this study, the MJ group, which performed the same multi-joint exercises as the MJS group, showed no change in hs-CRP. This may be because self-efficacy plays a role in reducing inflammation, as behavior change is a key factor. Dzerounian et al. [30] found that self-efficacy and health knowledge are linked to positive behavioral changes and increased physical activity in older adults, suggesting that health knowledge enhances self-efficacy, which in turn supports better health behaviors. Therefore, the increase in FPG and hs-CRP in the MJ group may reflect an insufficient adaptation to the exercise stimulus without the motivational reinforcement provided by self-efficacy theory.

This finding aligns with the results related to knowledge, self-efficacy, and outcome expectations, indicating that the MJS group experienced greater improvements than the MJ group following the interventions. This study highlights that individuals with high self-efficacy, combined with strong outcome expectations, develop increased self-confidence, which significantly influences behavior modification. As Bandura and Adams [31] suggested, individuals with high self-efficacy and positive outcome expectations are more likely to adopt and sustain health behavior changes. Consistent with previous research, strong self-efficacy and favorable outcome expectations encourage individuals to maintain regular exercise or physical activity [32]. Chu and Wang [33] found that outcome expectations for exercise are associated with perceived health and self-efficacy in older adults. Programs designed to enhance self-efficacy can effectively increase self-efficacy levels, leading to improvements in self-care activities [34, 35]. Additionally, understanding the stages of exercise behavior and exercise self-efficacy provides valuable insights for improving exercise adaptation and adherence, which, in turn, influences the overall healthcare of overweight or obesity older adults. Enhanced exercise adherence can have a significant impact on longevity, quality of life, and healthcare costs in the aging population [36].

These findings have important implications for designing exercise programs for overweight and obesity older adults. Incorporating psychological strategies such as self-efficacy theory could enhance the effectiveness of physical activity interventions, particularly for body composition and metabolic health. However, there are some limitations. One limitation of this study was the absence of follow-up assessments to evaluate the continuity of exercise behavior and self-efficacy after the intervention. It is recommended that future studies incorporate a 6-week follow-up period to examine the long-term sustainability of health-related outcomes and self-efficacy in overweight and obesity older adults. Additionally, the study lacked a control group performing only daily life activities, limiting the ability to attribute observed effects solely to the intervention. Furthermore, the gender distribution was imbalanced, with a lower proportion of male participants, which may limit the generalizability of the findings across sexes. Future studies should include follow-up assessments (e.g., at 6 weeks or longer) to examine the long-term effects on exercise behavior and self-efficacy. Including a true control group engaging only in daily activities would help clarify the intervention’s effectiveness. Moreover, ensuring balanced gender representation would improve the generalizability of the findings across sexes.

## Conclusion

Integrating self-efficacy theory with multi-joint exercise yields greater improvements in body composition, BMI, fat mass, and HbA1c levels compared to exercise alone. These findings underscore the importance of combining physical and psychological strategies to optimize health outcomes in overweight and obesity older adults. This approach may be applied in the development of community-based exercise programs, clinical rehabilitation protocols, and public health initiatives aimed at promoting long-term behavioral change, improving metabolic health, and enhancing overall quality of life in aging populations.

## Data Availability

All relevant data are within the paper and its Supporting Information files. Additional datasets will be made available in the OSF repository upon publication.

## Acknowledgments

The authors would like to express their sincere gratitude to all participants who took part in this study. We also extend our appreciation to the Senior Citizens’ Quality of Life Development Center, Nonthaburi City Municipality, for their kind support in providing the research site. Special thanks are due to the research assistants for their valuable contributions throughout the data collection process.

